# Pronounced State Level Disparities in Medicaid Prescribing of Buprenorphine for Opioid Use Disorder (2019-2020)

**DOI:** 10.1101/2022.09.28.22280256

**Authors:** Sydney R. Dana, Stephanie D. Nichols, Kenneth L. McCall, Brian J. Piper

## Abstract

**Objective:** To analyze buprenorphine prescription distribution across states in Medicaid patients during 2019-2020.

**Methods:** Buprenorphine prescriptions per Medicaid enrollee per state was calculated for 2019 and 2020. The totals of monoproduct buprenorphine were divided over the total of combination buprenorphine/naloxone in 2019 and 2020 to obtain the ratio of mono/combo. Data analysis was conducted with generic and brand name formulations of buprenorphine, and transmucosal buprenorphine/ naloxone combinations FDA-approved for OUD using Microsoft Excel. Formulations of buprenorphine indicated for pain were excluded. States outside 95% confidence intervals (mean +1.96*SD) were considered statistically significant.

**Results:** The overall change in buprenorphine prescribing between 2019 and 2020 was modest (+3.6%) but highly variable with > 10% increases in 17 states (Iowa = +100.5%, p < 0.05) but > 10% decrease in nine states (Alabama = −68.5%, p < 0.05). Total amount reimbursed in 2019 increased (+9.9%) to $1.42 billion in 2020. Branded formulations accounted for two-fifths (39.5%) of prescribing but over two-thirds (66.8%) of spending in 2020. There was a 50-fold difference in prescribing between the highest (Vermont = 219.5 prescription / 1,000 enrollees, p < 0.05) and lowest (Texas = 4.4) states in 2019. This disparity increased in 2020 resulting in a 278-fold difference.

**Conclusions:** The COVID-19 pandemic exacerbated state-level disparities in buprenorphine prescribing for OUD among Medicaid patients. Legislation on buprenorphine waivered providers and Medicaid expansion may explain the statistically significant percent changes in state buprenorphine prescriptions.

## Introduction

The number of prescription opioid overdoses rose 16.1% from 14,139 in 2019 to 16,416 in 2020 after steadily decreasing since 2016.^1^ Despite the clear mortality benefit of medications for opioid use disorder (MOUD), curiously, the majority of patients that have a non-fatal opioid overdose do not subsequently receive MOUD. ^2^ This post-overdose healthcare touch point represents an important opportunity to engage a patient in MOUD; unfortunately, the opportunity is often missed.

Before the COVID-19 pandemic, there were several barriers preventing patients from receiving MOUD buprenorphine, including challenges regarding transportation, child/eldercare, and lack of income, time, or other recourses. An important solution to many of these barriers, telemedicine, was prohibited as an access point for MOUD in 2008 by the Ryan Height Act which legislated that patients needed to be evaluated in person in order to be prescribed controlled substances, which includes the DEA Schedule C-III medication buprenorphine. ^3^ However, after the COVID-19 pandemic was declared a public health emergency in 2020, telemedicine as a method for comprehensive OUD treatment including buprenorphine therapy was possible. Telemedicine has increased access to buprenorphine, making obtaining treatment easier and more convenient for people who need it. ^3^

While buprenorphine and methadone are core MOUDs, use of buprenorphine in particular has been increasing over the past 2 decades. ^4,5^ Buprenorphine is a partial mu agonist with a high affinity for the mu receptor and a slow dissociation from the receptor, making it more difficult to displace. However, at typical OUD doses (e.g. 16 mg), buprenorphine occupies about 80% of mu receptors, leaving some unoccupied receptors that can be accessed for acute analgesic purposes when needed. ^6,7^ Further, as a partial mu agonist, buprenorphine has a reduced risk of nonmedical use and diversion, and a much lower risk of overdose than the full agonist methadone.^8^ Although some buprenorphine diversion is known to occur, ^9^ the majority of people who use buprenorphine outside of the medical system do so primarily to self-treat OUD (58% with first use and 71% with last use). ^10^ In contrast, getting high or altering mood was the primary reason for only 24% of people who use first used buprenorphine nonmedically and 19% who most recently used. ^10^ When given multiple selections for nonmedical use, the most common reasons were to prevent withdrawal (79%), maintain abstinence (67%), and self-wean off drugs (53%). ^11^ Of note, given the choice of yes/no with this multi-selection format, one-half (52%) of people also reported using buprenorphine to get high or alter mood, but only 4% indicated that it was their drug of choice. Together, these data suggest that lack of access to buprenorphine treatment is the primary reason for buprenorphine use outside the medical system and represents an important policy opportunity. ^11^

There is increasing appreciation that the buprenorphine receptor binding profile is more complex than simply as a mu partial agonist. Buprenorphine is active at all four of the opioid receptors, mu, kappa, delta, and nociception. ^12, 13^ Agonism of mu^1^ and mu^2^ receptors in the central nervous system causes euphoria, analgesia, nausea and vomiting, and respiratory depression. ^8^ Antagonism of the kappa receptor causes dysphoria. ^8^ Buprenorphine is a mu agonist and kappa and delta antagonist. It additionally has moderate affinity for the nociceptin receptor.Buprenorphine is metabolized in the liver by CYP450 3A4 to its active metabolite norbuprenorphine. ^12, 13^ This metabolite has a greater affinity for the nociceptin receptor and is partially responsible for the respiratory depression that may occur after buprenorphine administration. ^12, 13^ Respiratory depression is particularly a concern when buprenorphine is combined with benzodiazepines. ^14^ While the overall risk of overdose with buprenorphine alone is lower than full agonists, in a study of over 63,000 people on buprenorphine treatment, use of concomitant benzodiazepine tripled risk of fatal overdose and doubled risk of nonfatal overdose.^15^

These properties make buprenorphine different than most other opioids. Adding naloxone, an opioid antagonist, in combination with naloxone to treat OUD may reduce the potential for nonmedical use compared to methadone. Buprenorphine monoproduct is the FDA recommended MOUD for pregnant individuals, however building evidence illustrates the safety of combination product in this patient population. ^16^ In most other cases the combination product buprenorphine/naloxone is preferred. ^16^ Another contrasting feature of buprenorphine and methadone is weight gain; chronic MOUD with methadone caused body mass index elevations (2.2 to 5.4) and buprenorphine for OUD may be less likely to impact this. ^17^

To be able to prescribe buprenorphine for OUD, a provider must have an “X” DEA number and thus becomes a buprenorphine waivered prescriber. ^18^ Traditionally to be eligible, a provider must have completed specialized education, eight hours for physicians and 24hrs for physician assistants and nurse practitioners.^18^ This requirement recently changed making the educational component optional, however a prescriber must still apply for the X-waiver leaving a major access barrier intact. Astonishingly, over half (56.3%) of rural counties in 2017 lacked even one waivered provider. ^19^

Medicaid patients are prescribed buprenorphine mono- and combination products for OUD. To be eligible for Medicaid, a person must meet criteria that is decided by the United States Government through the Centers for Medicaid and Medicare Services. The Affordable Care Act established in March 2010 has amended the qualifications of eligibility for Medicaid insurance, resulting in Medicaid expansion in 39 states and territories including Washington DC as of June 2022. ^20^ People are eligible for Medicaid if they make at or below 138% of their state’s poverty level. ^21^ Some other examples of populations who are eligible include people who are pregnant people, children, and/ or people diagnosed with tuberculosis. ^22^

Most patients, including Medicaid patients, that receive buprenorphine are prescribed combination product (buprenorphine/naloxone) instead of mono product (buprenorphine). A prior study examined the state variations in buprenorphine prescribing to US Medicaid patients in 2011 through 2018. ^23^ Vermont had the highest rates of buprenorphine prescriptions among Medicaid patients, ^23^ and it was suspected that state will continue as the top buprenorphine prescribing nationally. Moving forward, quantification of the change and variations between 2019 and 2020 in buprenorphine prescribing among Medicaid enrollees is important to inform efforts to reverse escalating opioid overdoses and highlight opportunities for targeted public health initiatives.^1^

## Methods

### Procedures

Fifty-one states, included the District of Columbia, were the subject of this study. All national and state totals for buprenorphine prescriptions in Medicaid patients were obtained from [24]. The number of Medicaid patients in each state was obtained from Medicaid.gov as well. The mono buprenorphine and combo buprenorphine formulations that were primarily prescribed for OUD including generic buprenorphine and buprenorphine/naloxone and select brand names (Sublocade, Suboxone, Zubsolv, and Bunavail), and amount reimbursed were extracted. Using the NDC numbers provided with each prescription the FDA database was used to make the pain or OUD classification (Supplemental Table 1). All pain formulations buprenorphine and buprenorphine/naloxone were excluded in this study. Procedures were approved as exempt by the Geisinger IRB.

### Data-analysis

Ratios of buprenorphine prescriptions per 1,000 Medicaid enrollees per state were calculated for 2019 and 2020. Data analysis was conducted with mono product and combo, generic and brand, and amount reimbursed of OUD buprenorphine using Microsoft Excel. The 95% confidence intervals were calculated for the 2019, 2020 and percent change intervals using the Excel equation (=CONFIDENCE(a,stdev,n)). Statistical significance and correlation coefficients were completed with Excel. Statistical significance was calculated using the equation (=MEAN +/-1.96* STDEV.P) and correlation coefficients were calculated using (=correl(a:a,b:b)). Fold changes between states were calculated by dividing the highest value by the lowest value in Figures 1 and 2, respectively. Mono and combo product and brand/ generic classifications were gained by searching the NDC numbers given with each prescription of buprenorphine on this website ndclist.com. ^25^ All heatmaps were made using USA-Heat-Map-Someka-Excel-V15 from someka.net. All graphs were prepared with Graph Pad Prism. Those distributions were visualized on heat maps and graphs using Someka Software and Graph Pad Prism, respectively.

**Figure 1.**
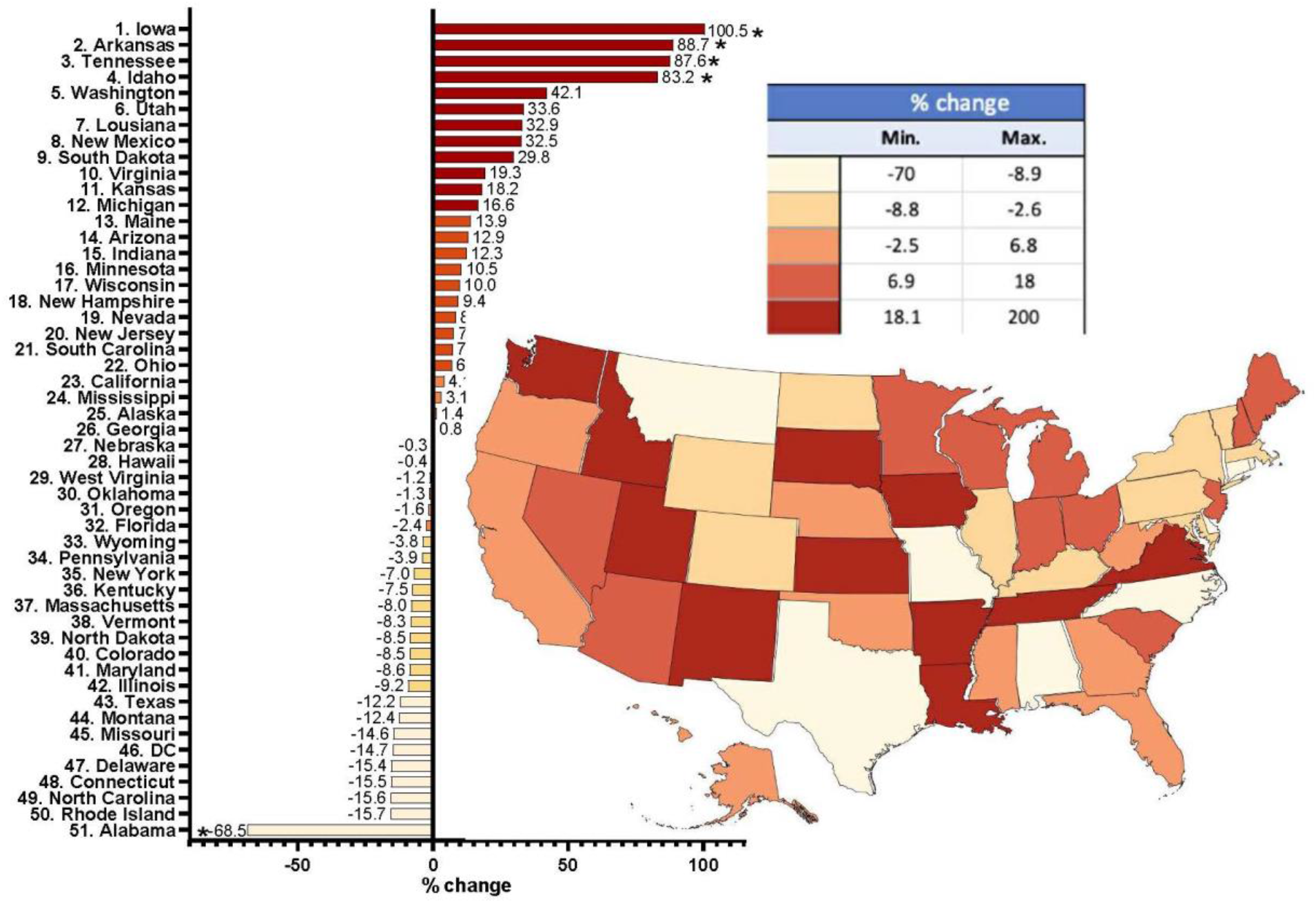
The percent change in buprenorphine prescribing for Opioid Use Disorder to Medicaid patients between 2019 and 2020. The states that had statistically significant increases were Iowa (100.5), Arkansas (88.7), Tennessee (87.6) and Idaho (83.2). Alabama (−68.5) has a statically significant decrease between 2019 and 2020. 95% CI [-49.1, 65.9].

**Figure 2.**
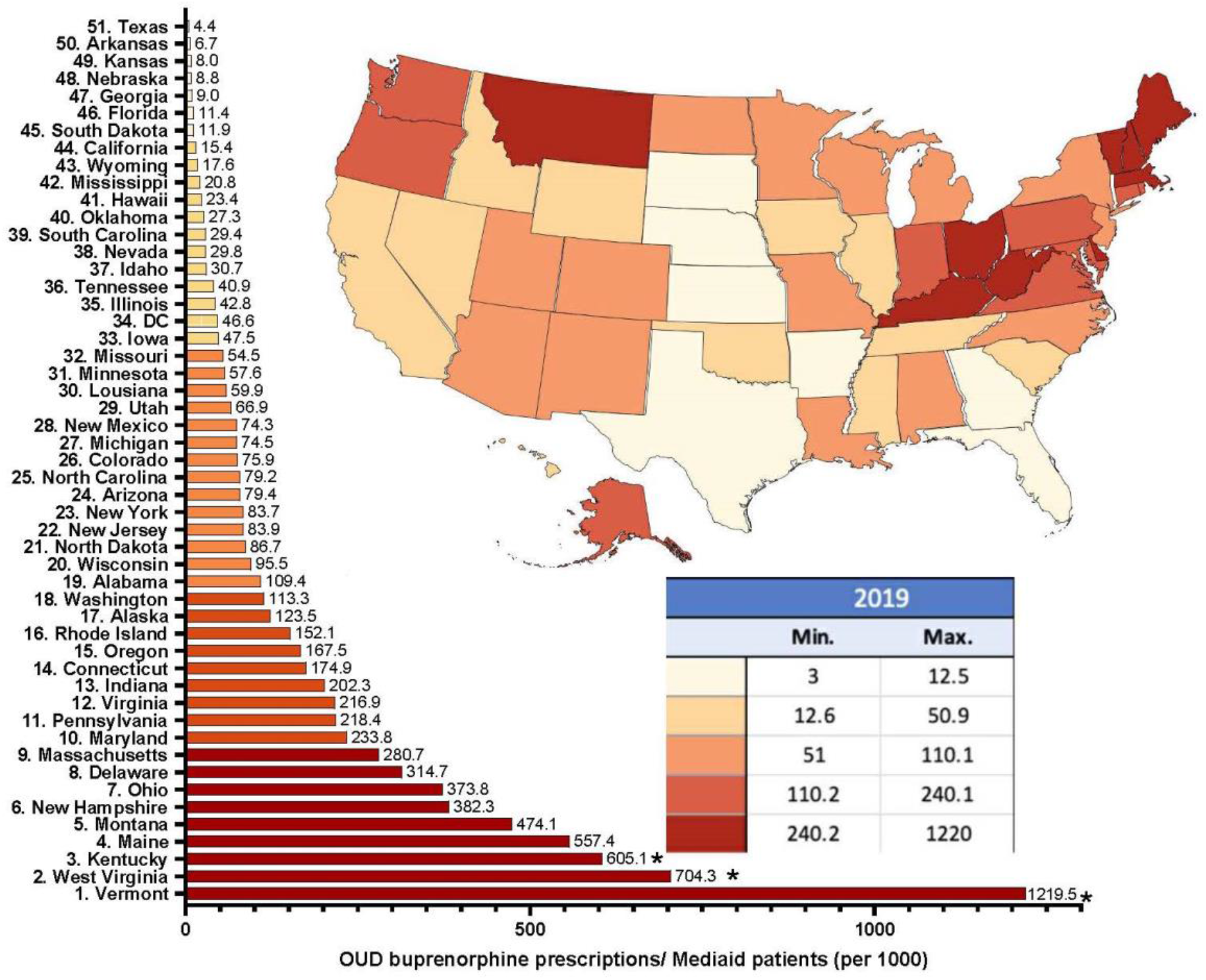
Buprenorphine prescriptions for Opioid Use Disorder in 2019 per 1,000 Medicaid enrollees. Three states were statistically significant (*p < .05) and outside the 95% confidence interval [-275.2, 570.2].

## Results

This study examined the number of buprenorphine and buprenorphine/naloxone prescriptions for OUD in Medicaid patients across the 51 states in the US and showed a slight (+3.64%), but regionally dependent, elevation between 2019 and 2020. Four states with greater than 80% increases, Iowa, Arkansas, Tennessee, and Idaho, were each significant relevant to the average state (8.4%). Almost twice as many states (17) had a > 10% elevation as had as underwent a decline of the same magnitude (9, Figure 1).

Further examination revealed a pronounced (49.9) fold difference between the highest and lowest states in 2019 (Figure 2). Except for Montana, twelve of the thirteen highest prescribing states were east of the Mississippi River with clusters in New England (ME, NH, VT, MA) and Appalachia (WV, KY, OH).

The state level differences became even larger with a 286.8-fold difference between the highest (VT = 1,118.5 prescriptions / 1,000) and lowest (TX = 3.9 / 1,000) states in 2020 (Figure 3). Besides Montana, twelve of the thirteen highest states were again located east of the Mississippi River with clusters in New England (ME, NH, VT, MA) and Appalachia (WV, VA, KY, OH).

**Figure 3.**
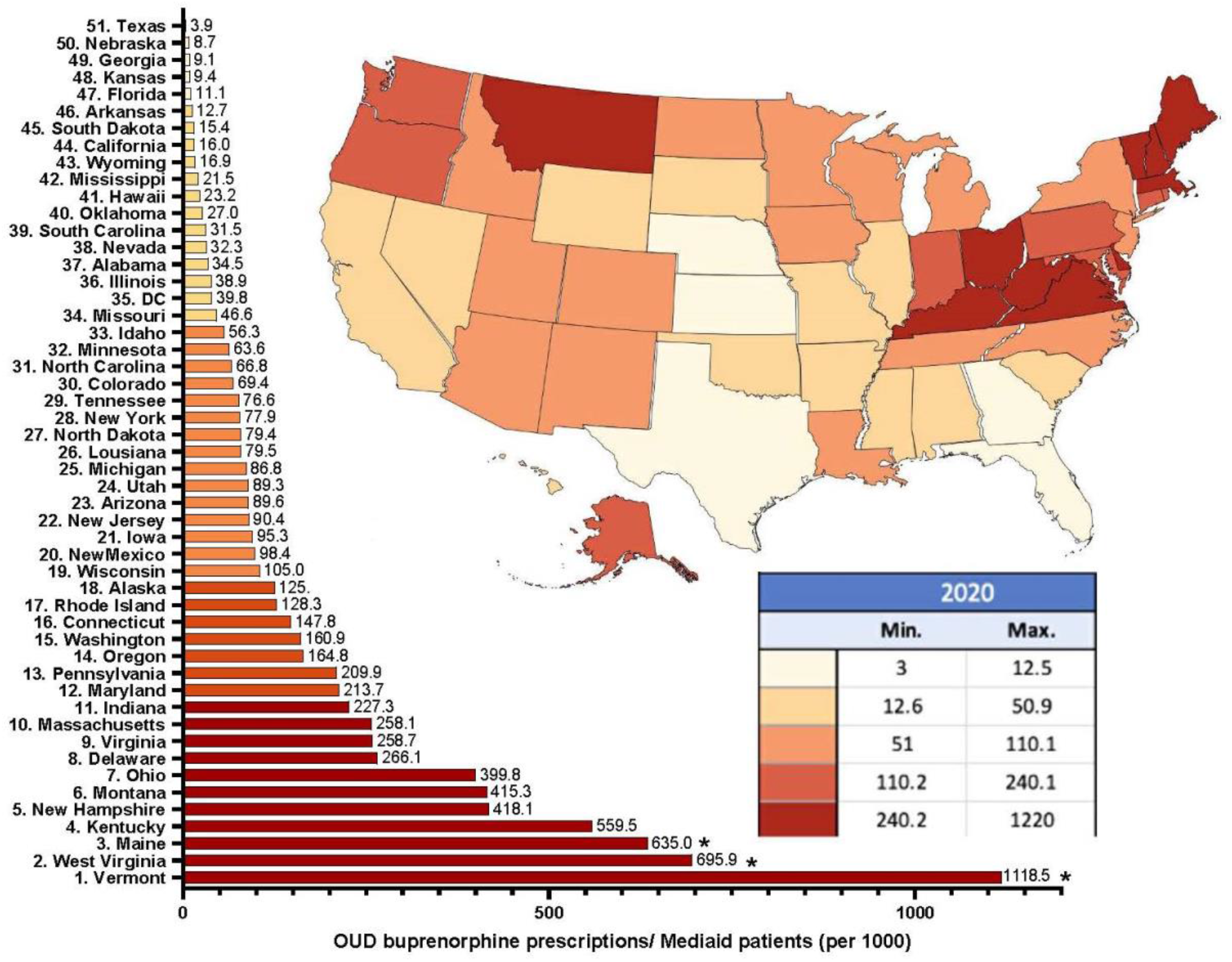
Buprenorphine prescriptions in 2020 for Opioid Use Disorder per 1,000 Medicaid enrollees. Three states were outside (*p < .05) the 95% confidence interval [-255.5, 570.2].

A correlation coefficient was calculated between prescriptions in 2019 and 2020 and showed a strong association (r(49) =0.991, p < .001).

Correlation coefficients were calculated between the ratio of combo/ mono product of buprenorphine prescriptions and the total OUD buprenorphine prescriptions per Medicaid enrollee for each year. In 2019, the correlation coefficient was r(49)=0.476 and in 2020 it was r(49)=0.516, which were significant.

In 2019, out of the total prescriptions of buprenorphine mono and combo products, 3.98 million prescriptions were brand name and 3.90 million were generic buprenorphine prescription (49.5%). In 2020 the split between brand name and generic prescriptions was not as evenly split.

The brand name prescriptions made up 3.4 million of the 8.5 million (39.4%) of the total buprenorphine prescriptions.

The total amount reimbursed for buprenorphine OUD prescriptions in 2019 was just shy of $1.29 billion and increased slightly (+9.9%) to $1.42 billion in 2020. Branded formulations accounted for 72.6% of spending in 2019 and 66.8% in 2020.

## Discussion

Despite the national escalation in drug overdoses, ^1^ and the well-established role of buprenorphine in reducing opioid overdoses, ^4,5^ we identified relatively small changes (<4%) nationally in buprenorphine prescribing to Medicaid patients during the first year of the COVID-19 pandemic. Further, and of great concern given the climbing overdose rates, half of states (25) decreased their prescribing.

This report found that Vermont stayed the highest buprenorphine prescriber in 2019 and 2020, Iowa, Arkansas, Tennessee, Idaho and Alabama all had statistically significant changes in buprenorphine prescriptions between 2019 and 2020, and that Washington state had a pronounced surge of combination buprenorphine prescriptions in Medicaid patients in 2019. In addition to federal policy changes there were state specific legislation and policy changes that may account for the tremendous differences between states. ^26^ There are federal regulations but often states also make regulations that buprenorphine waivered prescribers must follow.

Vermont, West Virginia, Kentucky, Maine, Iowa, Arkansas, Tennessee, and Idaho were among the states that had statistically significant results (Figures 1–3).

Vermont has been a leader since 2014 in paving the way in buprenorphine prescribing with their hub and spoke model.^27^ This model has put Vermont at the top of buprenorphine prescriptions since then with little variation according to a report that tracked the buprenorphine prescriptions in US Medicaid patients in 2011 through 2018. It has led to a 64% increase in buprenorphine waivered prescribers from 2014 to 2019. ^27^ In 2019 and 2020, Vermont was statistically significant in having the most buprenorphine prescriptions per 1,000 Medicaid patients. The hubs in the model is where people receiving MOUD for OUD can receive intensive treatment with methadone, buprenorphine and naloxone and the spokes are maintenance centers where people can receive continuing care for OUD with buprenorphine and naloxone. ^27^ Not only did Vermont implement this model in 2014 but they also underwent a Medicaid expansion. ^20^ This allowed for more patients to be covered under Medicaid insurance and therefore allowed more people to receive treatment with buprenorphine for OUD. To add this also effected Idaho as they also underwent a Medicaid expansion in 2020 which could explain the percent increase in Figure 1.^20^

Second to Vermont in Figures 2 and 3 was West Virginia. In 2018 there was discussion for West Virginia to adopt a modified version of the Hub and Spoke model posed first by Vermont. In this adoption of the model, five hubs were established throughout West Virginia in hopes of increasing the capacity for OUD treatment with buprenorphine. In a study published there was statistically significant increases in buprenorphine distribution following the establishment of the five hubs. ^28^ Additionally, like Vermont, West Virginia also expanded Medicaid in 2014. ^20^

Kentucky and Maine followed Vermont and West Virginia in both 2019 and 2020, respectively. Both states had a Medicaid expansion, Kentucky in 2014 and Maine in 2019. This Medicaid expansion in Maine could have contributed to the rise in prescriptions in 2020 making the number of prescriptions statistically significant. ^20^ In Kentucky there is legislation that removes prior authorization for combination buprenorphine prescriptions in 2018. ^29,30^ In 2019 and 2020 the ratio mono/combo buprenorphine therapies Kentucky fell in the bottom 20% of all 51 states, meaning there was a large amount more combination buprenorphine prescriptions than mono product therapies. Which can be explained by the prior authorization wave in combination therapies only. ^29, 30^

In Figure 1, the five states that showed statistically significant percent changes were Iowa, Arkansas, Tennessee, Idaho, and Alabama. Iowa and Arkansas both had changes in legislation that removed prior authorization for buprenorphine prescriptions. This allowed for easier access to buprenorphine prescriptions. In Arkansas this took place in the beginning of 2020 and in Iowa it happened in the middle of 2019. ^31,32^

Even though Arkansas had the second largest percent increase between 2019 and 2020, the state has low numbers overall when looking at Figures 2 and 3. However, the increase in percent change can be attributed to the Arkansas Medicaid Pharmacy removing preauthorization for buprenorphine prescriptions in early 2020. ^32^

In 2020 Tennessee law makers made legislation that allowed nurse practitioners and physician assistants to be eligible to be buprenorphine waivered prescribers. ^33^ This expanded the care for OUD into rural communities and areas in Tennessee where people may have lacked accessibility to treatment.

Alabama was the only state in Figure 1 to have a statistically significant percent decrease between 2019 and 2020. This distribution of buprenorphine prescriptions could be due the controversy in the state regarding the medical committee formed in May 2019 tasked with making decisions on how to treat OUD using MOUD. This increased the number of restrictions on buprenorphine prescriptions in hopes of stopping opioid addiction. ^34^ The laws made by this committee included a regulation that prescribers had to see the patient for 12 months prior to prescribing opioids. ^35^

States in the bottom five of prescriptions of buprenorphine in 2019 and 2020 were Texas, Georgia, and Kansas. These states have not adopted Medicaid expansion, and therefore have those low prescription numbers to Medicaid patients. ^20^ Nebraska was also consistently in the bottom five of buprenorphine prescriptions however the state has implemented Medicaid expansion in Oct of 2020, but that might not have been enough time to see a rise in prescriptions^20^.

The wide fold change in both 2019 and 2020 can be attributed to the hub and spoke model that Vermont has adopted that has been successful in the distribution of buprenorphine and the lack of Medicaid expansion in Texas.

The heatmaps showing buprenorphine prescribing by state shows a moderate correspondence with the drug overdose rates (highest in WV, KY, OH, TN, and PA; lowest in ND, SD, NB, KS, and TX). ^36^ However, this association should be viewed carefully as the lowest states in the central US (ND, SD, NB, KS, TX) have a coroner driven process which is known to face challenges in accurate death determinations.^37,38^ Based on other reports,^39^ we are also skeptical that states with the lowest buprenorphine prescribing might have compensated with elevated methadone for OUD prescribing. Future study will be necessary to determine the extent that persistent logistical barriers continue to impair MOUD availability,^40^ particularly given the sizable state level variation identified.

Since there were statically significant correlation coefficients between the number of prescriptions per 1000 Medicaid patients and the combo/ mono product for 2019 and 2020, the increase in total prescriptions was correlated to the increase in combo product prescriptions.

In both years, 2019 and 2020, North Dakota and West Virginia had statistically high ratios of combo/ mono product seen in Figures 4 and 5. West Virginia, as discussed earlier, has been steadily increasing buprenorphine prescriptions to try and combat the growing number of people diagnosed with OUD. ^20,27^ It is suspected that telehealth had a large part in the 105% increase in combo/mono buprenorphine prescriptions seen in Figures 4 and 5 in North Dakota. ^41^

**Figure 4.**
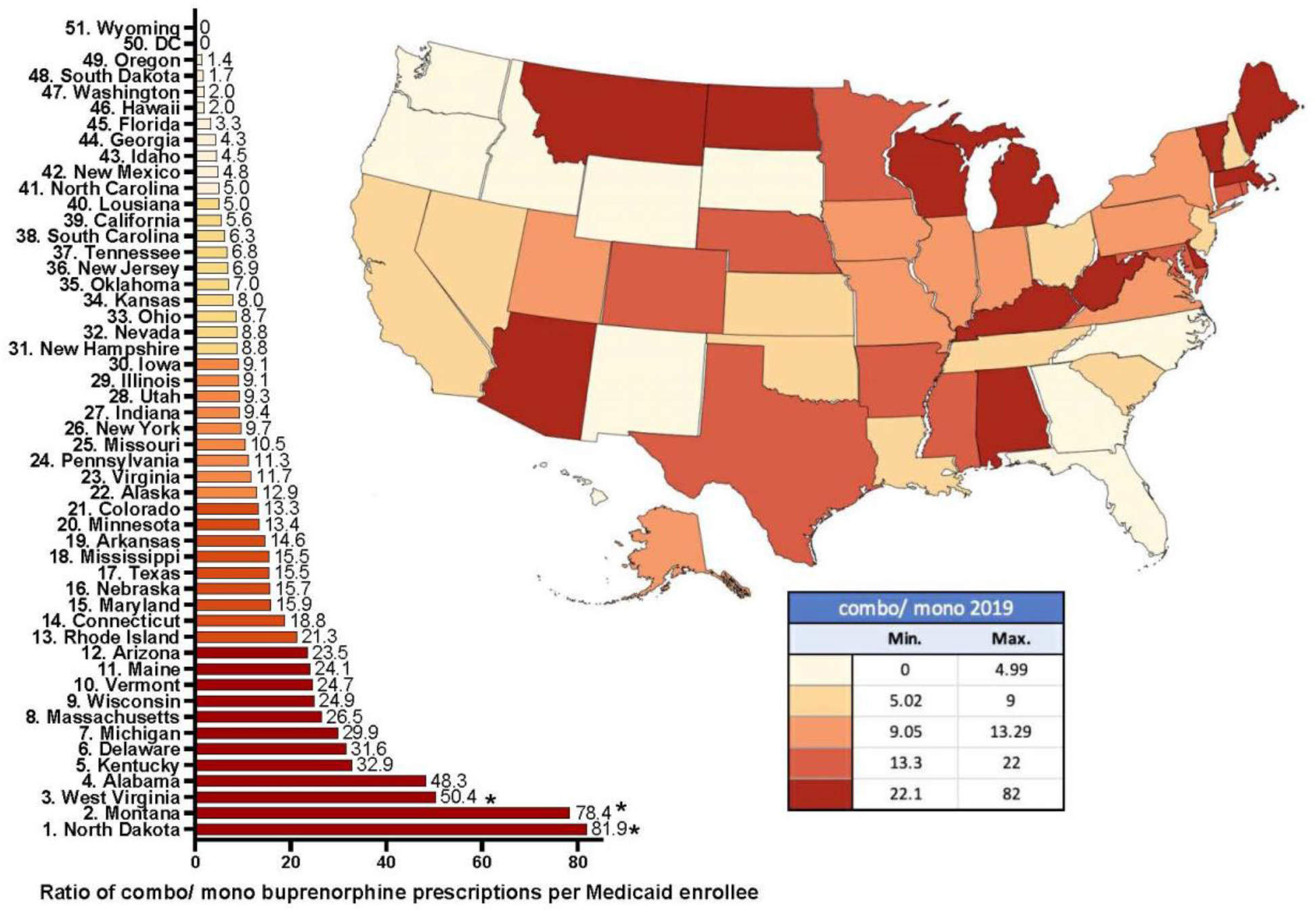
Ratio of combination to mono product buprenorphine prescriptions per Medicaid enrollee in each state in 2019. North Dakota (81.9), Montana (78.4), and West Virginia (50.4) were all statistically high (*p < .05) and outside the 95% CI [-17.4, 49.2]. Wyoming and the District of Columbia (DC) had zero mono product prescriptions.

**Figure 5.**
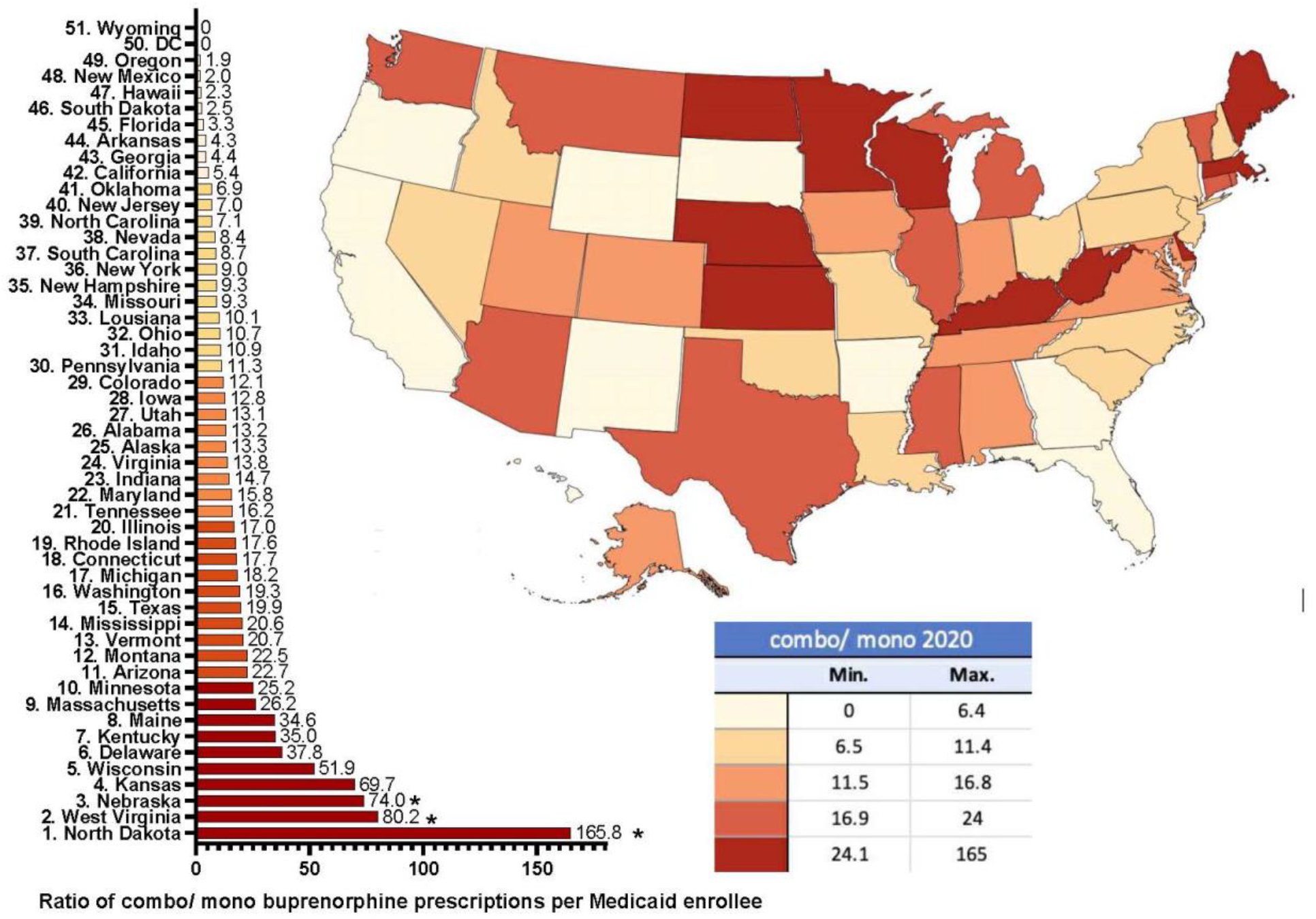
Ratio of combination to mono product buprenorphine prescriptions per Medicaid enrollee in each state during 2020. North Dakota (165.8), West Virginia (80.2), and Nebraska (74.0) were all statistically high (*p < .05) and outside the 95% CI [-32.3, 73.4]. Wyoming and the District of Columbia (DC) had zero mono product prescriptions.

In June of 2018 Dr. Reddy Laboratories, a multinational pharmaceutical company, received FDA approval for a generic Suboxone sublingual film product. However, later in that same year they were blocked from production of their product by a temporary restraining order filed by Indivior UK Unlimited. ^42^ The court of appeals in December 2018 ruled in favor of Dr. Reddy Laboratories and they began generic Suboxone production. ^42^ This powerful lawsuit could contribute to the differences seen between brand and generic product ratios in 2019 and 2020. ^42^

Overall, despite federal policy changes to enable greater use of telemedicine, ^41^ the modest change in prescribing nationally (3.6%), coupled with the substantial state level differences for Medicaid patients in the face of escalating illicit and prescription opioid overdoses is concerning. ^1, 2^ Due to the stigma associated with addiction, it is extremely difficult to even quantify how many OUD patients are in the US with estimates as high as 7.6 million. ^43^ Clearly, further efforts to increase MOUD availability and reduce barriers are pressing public health needs.

Looking at overdose rates in these states between 2019 and 2020 and years later would be an indication that the legislation made between 2019 and 2020 had an impact in treatment of people with OUD. The distributions between buprenorphine prescriptions and prescription overdoses would give additional insight into where treatment could improve and how. Another way this study could be expanded upon is comparing state-by-state differences in route of administration. Additionally, how the median dose of buprenorphine per prescription compared to overdose deaths in each state.

## Limitations

Some caveats and future directions are noteworthy. First, further study should quantify the number of buprenorphine waivered prescribers including physicians, ^44^ and advanced practice providers in each state for 2019 and 2020 to determine if the aforementioned legislative implementations had a direct impact on the changes on buprenorphine prescriptions in Medicaid patients. ^19^ Second, even though the NDC indicates that a prescription was used for OUD vs pain, we only examined those identified as for OUD (Supplemental Table 1). We are aware that some off-label use is likely to have occurred although we presume that this level was relatively homogenous across states. Third, methadone by weight has almost three-fold greater distribution than does buprenorphine in the US although this gap may be narrowing.^45^ Future investigations should examine whether there are similar state level disparities in methadone prescribing to Medicaid and Medicare patients.

## Conclusion

In conclusion, looking at the years 2019 and 2020 there were gross, but modest, changes in buprenorphine prescriptions to Medicaid patients being treated for OUD. As predicted Vermont remained the highest buprenorphine prescriber nationally in 2019 and 2020. As discussed, policy can significantly influence buprenorphine prescribing and thus policies aimed at increasing access to buprenorphine, including the removal of the X-waiver requirement, should be considered. Increasing the availability to medically treat patients with OUD is a pressing public health need.

## Data Availability

All data produced in the present study are available upon reasonable request to the authors.

## Acknowledgements

The contributions of Lauren Donahoe, Madison Frank, Madison Hurst, Caitlin Guinee, Joanna Bernatowicz, and Iris Johnston are recognized.

---

**Supplemental Table 1.**
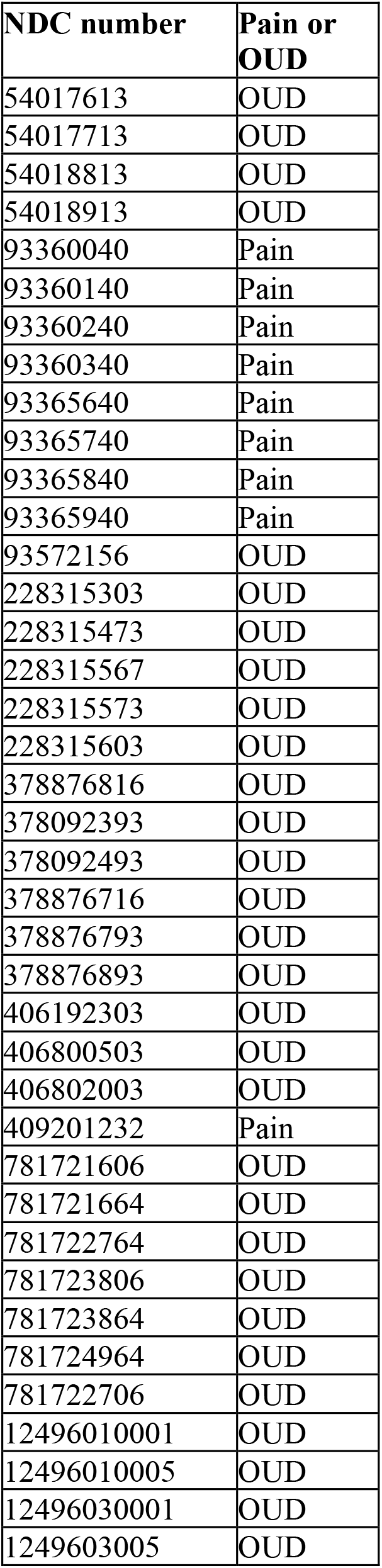
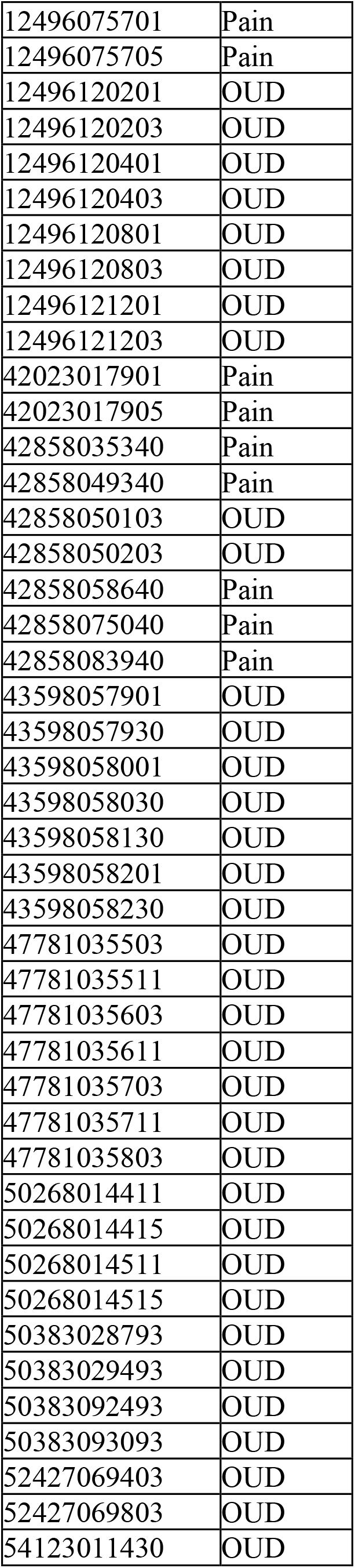
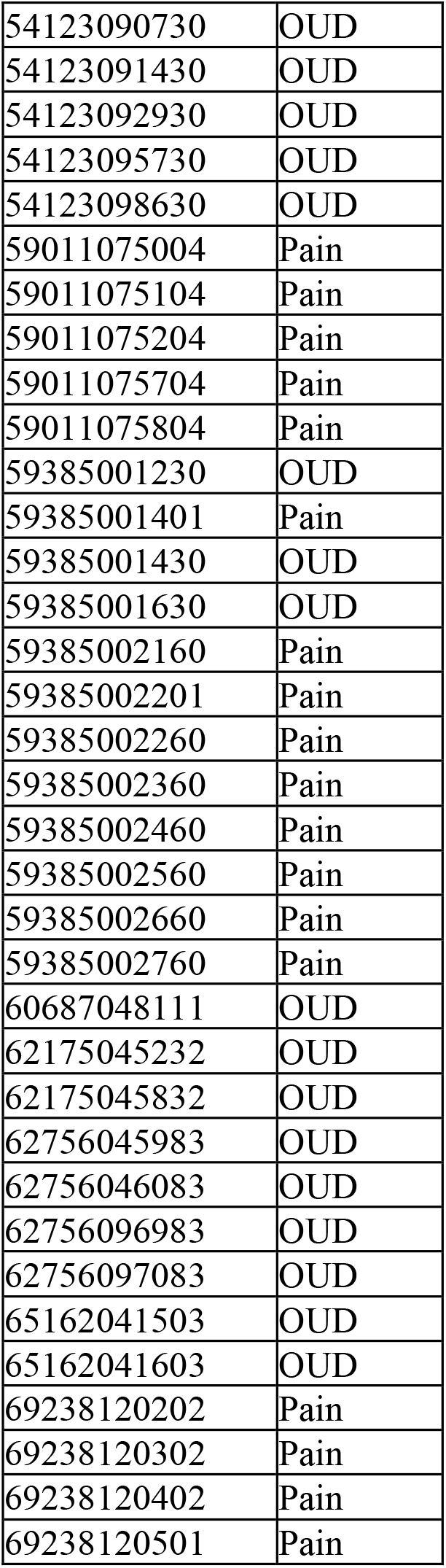
National Drug Code (NDC) numbers and their respective Pain or Opioid Use Disorder (OUD) classifications.

**Supplemental Table 2.** Brand name formulations of buprenorphine claims from 2019 to 2020 among Medicaid patients.

| Year | Prescriptions<br>Zubsolv | Prescriptions<br>Suboxone | Prescriptions<br>Sublocade |
| --- | --- | --- | --- |
| 2019 | 151,611 | 3,706,879 | 27,089 |
| 2020 | 133,748 | 3,068,577 | 59,567 |
| % Change | -11.8% | -17.2% | 119.9% |

